# Impact Of an Immune Modulator Mycobacterium-w On Adaptive Natural Killer Cells and Protection Against COVID-19

**DOI:** 10.1101/2021.12.14.21267696

**Authors:** Sarita Rani Jaiswal, Jaganath Arunachalam, Ashraf Saifullah, Rohit Lakhchaura, Dhanir Tailor, Anupama Mehta, Gitali Bhagawati, Hemamalini Aiyer, Bakulesh Khamar, Sanjay V. Malhotra, Suparno Chakrabarti

## Abstract

The kinetics of NKG2C^+^ adaptive natural killer (ANK) cells and NKG2A^+^inhibitory NK (iNK) cells with respect to the incidence of SARS-CoV-2 infection were studied for 6 months in a cohort of health-care workers following administration of heat killed *Mycobacterium w* (Mw group) in comparison to a control group. In both groups, COVID-19 correlated with a lower NKG2C^+^ANK cells at baseline. There was a significant upregulation of NKG2C expression and IFN-γ release in Mw group (p=0.0009), particularly in those with lower baseline NKG2C expression, along with downregulation of iNK cells (p<0.0001). This translated to a significant reduction in incidence and severity of COVID-19 in the Mw group (IRR-0.15, p=0.0004). RNA-seq analysis at 6 months showed an upregulation of ANK pathway genes and an enhanced ANK mediated ADCC signature. Thus, Mw was observed to have a salutary impact on the ANK cell profile and a long-term upregulation of ANK-ADCC pathways, which could have provided protection against COVID-19 in a non-immune high-risk population.

## INTRODUCTION

The rapid explosion of the novel coronavirus, SARS-CoV-2, in early 2020, across the globe overwhelmed even the most prepared health infrastructures[1] and exposed the healthcare workers to an unforeseen situation, where they remained at the greatest risk of exposure to the highest viral load, in the absence of a prevention or a cure. Despite a very high incidence of infections, witnessed in the Indian population as well, there was a surprising sparing of the urban slum-dwellers[2]. We hypothesized the role of a bolstered innate immune system secondary to chronic pathogen exposure as a plausible reason for this phenomenon. We reasoned that due to ubiquitous exposure to cytomegalovirus (CMV) in early childhood, followed by exposure to a multitude of pathogens subsequently, there might be stronger repertoire of NKG2C expressing adaptive natural killer (ANK) cells in this population [3].

Early CD56^bright^ NK cells have very high expression of NKG2A, which functions as an inhibitory check-point in the process of functional maturation of NK cells[4]. Both NKG2C and NKG2A bind to the same ligand, HLA-E, but the latter binds with several-fold greater affinity compared to NKG2C [5]. Unlike somatic mutations witnessed in adaptive immune cells to produce precise and clonal antigen specificity, NK cells express a plethora of germline encoded activating and inhibitory receptors. The regulation of NK cell function, which is termed as ‘licensing’, occurs with stochastic expression of killer-immunoglobulin-like receptors (KIR), which bind to self-HLA-Class 1 antigens with biallelic specificity[6]. Expression of KIRs for which appropriate self-HLA antigens exist enable continued inhibition of NK cells preventing autologous cytotoxicity. Hence, NKG2A^+^inhibitory NK cells (iNK) are key to the prevention of autoreactivity of NK cells, prior to the KIR-driven process of licensing. In a subset of mature and licensed NK cells, exposure to CMV leads to the expression of a C-lectin type activating receptor, NKG2C, which is encoded by the KLRC2 gene[7]. These cells are characterized by upregulation of NKG2C and downregulation of the inhibitory counterpart, NKG2A[8]. This subset of NK cells, now called NKG2C^+^ANK cells, exhibits the classic adaptive features, such as, clonal expansion, persistence and recall memory more akin to memory cytotoxic T cells, than canonical NK cells[9].

While the major subset of ANK cells express NKG2C, which is the defining phenotype, several myeloid (FCεRG, PLZF) and B lymphoid (SYK, EAT-2) genes are downregulated, and certain T lymphoid genes (CD3ζ, BCL11B) are upregulated in ANK cells[10]. The alterations in these gene expressions in ANK cells favour an augmentation of antibody-dependent cellular cytoxicity (ADCC). In a small subset of ANK cells, the adaptive functions might be demonstrable with epigenetic distribution of myeloid and lymphoid associated genes as described above, even without upregulation of NKG2C expression[10; 11]. Hence, for the sake of clarity, the ANK cells described in this study are NKG2C^+^ ANK cells.

In the context of haploidentical hematopoietic cell transplantation (HCT), NKG2C^+^ANK cells were found to afford protection, not only against leukemia, but also a range of viral infections, other than CMV. It was suggested that high NKG2C^+^ ANK cells was essential in maintaining a non-inflammatory milieu without compromising anti-tumor effect post-HCT[12; 13; 14; 15]. Based on these seminal findings along with the existing evidence that certain natural infections, such as Hantavirus, as well as Influenza and BCG vaccinations are capable of upregulating NKG2C^+^ANK cells in CMV-exposed populations[16; 17; 18], we hypothesized that a novel heat killed *Mycobacterium w* (Mw), also known as *Mycobacterium indicus pranii*, an approved immunomodulator in India [19], might upregulate NKG2C^+^ANK cells and offer protection against SARS-CoV-2 (Severe Acute Respiratory Syndrome Coronavirus-2) infections in the process. We studied the impact of Mw on the incidence of COVID-19 (Corona Virus Disease 2019 caused by SARS-CoV-2) in a cohort-control study in front line health care workers and its impact on the kinetics and repertoire of NKG2C^+^ANK cells in the context of KLRC2 genotype.

## MATERIALS AND METHODS

In a single-center non-randomised cohort control study, 50 HCWs from a single department in the hospital were administered 0.1 ml *Mw* (Sepsivac, Cadila Pharmaceuticals, India) intradermally in each arm on day 1 of the study (Mw group) and 50 randomly selected HCWs from the rest of the institution were enrolled in a Control group. In addition, in the Mw group, those without any local site reaction, who consented for second dose, were administered an additional dose of 0.1 ml Mw, 30 days after the first dose. The observation period of the study was 15 days after administration of the first dose of Mw to 180 days (September 2020 to February 2021).

All HCW included in the study had nasopharyngeal swab evaluated for SARS-CoV-2 by reverse transcriptase-polymerase chain reaction (RT-PCR), on development of symptoms suggestive of COVID-19 or following contact with a patient, HCW or a family member in contact with COVID-19 patient. Body temperature, pulse rate, oxygen saturation and self-reporting of symptoms was evaluated before and after each working day[20]. COVID-19 was diagnosed and its severity was graded as per established criteria. The duration of observation was 6 months from September 2020 to February 2021. In addition, blood was collected, at baseline, days 30, 60, and 100 from all the subjects, for evaluating the kinetics of NKG2C^+^ANK cells and T cells. All subjects of this study also underwent evaluation for CMV status (seropositive or seronegative).

All subjects provided written informed consent for participating in the study. The study was approved by institutional ethics committee and registered with Clinical Trials Registry of India (CTRI/2020/10/028326).

### Real Time Reverse Transcription Polymerase Chain Reaction (RT-PCR) for Detection of SARS-CoV-2

All Covid tests were done by Truenat Real Time Reverse Transcriptase Polymerase chain reaction (RT-PCR) test. Nasopharyngeal swabs were collected using standard nylon flocked swab and inserted into the viral transport medium (VTM) provided from the same company (Molbio diagnostics Pvt. Ltd. Goa, India). Samples were transported immediately to the laboratory maintaining proper temperature and processed as per manufacturer’s guidelines. [Truenat Beta CoV Chip-based Real time PCR test for Beta Coronavirus, Molbio diagnostics Pvt. Ltd. Goa, India]. The target sequence for this assay is E gene of Sarbeco virus and human RNaseP (internal positive control). Confirmatory genes were RdRP gene and ORF1A gene.

### Immunological monitoring

Peripheral blood mononuclear cells (PBMC) were isolated from whole blood samples, by density gradient centrifugations using HiSep™ LSM 1077 media. For surface staining, 0.5 × 10^6^ cells were washed with phosphate-buffered saline (PBS) and stained with the following antibodies which were used for phenotypic analysis: CD3(APC-H7, SK-7) CD16 (PE-Cy7, B73.1), CD56 (APC R700, NCAM16.2), CD57 (BV605, NK-1), NKG2A (PE-Cy7, Z199), CD4 (APC-H7), CD8 (Per-CP Cy), CD45RA (FITC), CD45RO (BV605), from BD Biosciences, (San Jose, CA) and NKG2C (PE, REA205) from Miltenyi Biotec, Germany. Cells were then incubated for 30 minutes. Viability was assessed with 7-AAD viability dye (Beckman Coulter). For intracellular staining, cells were stained for IFN-gamma using monoclonal antibodies for interferon-gamma (IFN-γ) (4S.B3) and perforin (Alexa647, DG9) (BD Biosciences) after fixation and permeabilization with appropriate buffer (BD Biosciences and e-biosciences, San Diego, CA, USA). Flow Cytometry was performed using 10 colour flow cytometry (BD FACS Lyrics) and the flow cytometry data was analyzed using FlowJo software (v10.6.2, FlowJo). Unstained, single stained (one antibody/sample) as well as fluorescence-minus-one (FMO) samples were used as controls for the acquisition as well as the subsequent analysis. Statistical divergences were determined by the GraphPad Prism software.

The gating strategy has been described earlier[15]. NKG2C^+^ANK cells were defined as CD56^dim^NKG2C^+^NKG2A^-^CD57^+^ subset of NK cells. NKG2C/NKG2A ratio was calculated as relative % of NKG2C^+^NKG2A^-^ ANK cells / NKG2C^-^NKG2A^+^ iNK cells (Fig. S4).

### KLRC2 (NKG2C) genotyping

KLRC2 gene encodes for NKG2C and is located in chromosome 12p13. For categorizing subjects KLRC2 wildtype homozygous (*Wt*^*+*^*/ Wt*^+^), KLRC2 deletion homogyzous *(Del*^*+*^*/ Del*^*+*^) and KLRC2 heterozygous (*Wt*^*+*^*/ Del*^*+*^), PCR-based KLRC2 genotyping was carried out. DNA was isolated from the peripheral blood using QIAGEN QIAamp@ DNA blood mini kit method (Hilden, Germany). PCR amplification was carried out with forward and reverse primer sequences as previously described[21]. PCR amplification was carried out in 20μl volume, containing 1x PCR master mix which has premixed taq polymerase, dNTPs, PCR buffer (Thermo fisher scientific, Waltham, MA 02451, United States), 1.65 pmol forward primers and reverse primers for KLRC2 deletion and wild type KLRC2 genes (see Table S2), 100pg-1μg genomic DNA. Amplification was performed using a T100 thermal cycler (Bio-Rad, Hercules, CA). Cycling temperature profiles were adopted from a previous study[21] with minor modifications.

Briefly, the reaction mixture was subjected to one cycle of denaturation at 95^°^C for 10 min followed immediately by 29 cycles of 95^°^C for 20 s, 50^°^C for 30 s, 72 ^°^C for 40 s; and a final extension at 72^°^C for 10 min before cooling to 4^°^C. A non-template control was included in each batch of PCR reactions. PCR products were identified by running the entire PCR product on a 2% agarose gel for 60 minutes at 70 Volts. The size of the amplicons was determined by comparison against the migration of a 100-bp DNA ladder (GeneDireX, Inc, Taoyuan County, Taiwan). Agarose gels were visualized and documented using the Gel doc XR+ gel documentation system (Bio-Rad, Hercules, CA). An illustration of the PCR assay is shown in supplementary figure (Fig. S5).

### RNA-seq Analysis

This was carried out on both Mw and Control group at 6 months following exposure to Mw. Four subjects from each group without COVID-19 were selected for the study.

### RNA isolation using Trizol method and polyA RNA selection

Peripheral blood mononuclear cells (PBMC) were isolated from whole blood samples by density gradient centrifugations using HiSep™ LSM 1077 media (Himedia, Mumbai, India). 5 × 10^6^ PBMCs were used for RNA isolation using the Trizol method and followed by DNase treatment for RNA purification. Purified RNA was quantified using a Qubit 4.0 fluorometer. Take 5μg of total RNA used for the polyA RNA selection using NEB NEXT oligo d(T)25 beads (NEB, MA, USA). PolyA RNA was quantified and integrity assessed using Qubit 4.0 fluorometer (Invitrogen, Waltham, Massachusetts, United States) according to manufacturer instructions.

### cDNA library preparation and whole transcriptome sequencing using MinION 2.0-Oxford Nanopore Technologies (ONT)

For Direct cDNA Native Barcoding Sequencing (SQK-DCS109 with EXP-NBD104, oxford nanopore technologies, Oxford, UK), 100ng of polyA RNA was used for the library preparation. Using H minus Reverse Transcriptase (Invitrogen, Waltham, Massachusetts, United States), complementary DNA (cDNA) was synthesised, followed by RNA degradation and second-strand synthesis of cDNA. Double-stranded cDNA was used for the end preparation, followed by native barcoding and adaptor ligation (all steps were followed, according to the manufacturer’s instructions). Ligated cDNA was loaded on the flow cell (R9) in MinION Libraries were sequenced specifying 72 hours on the ONT MinION using R9.4.1 flow cells and MinKNOW (v21.06.10, Microsoft Windows OS based) to generate FAST5 files. FAST5 files were base-called with CPU based Guppy basecaller (v.5.0.11) (ONT) to create summary text files and FASTQ files of the reads for further downstream analysis.

### Sample pre-processing and Quality assessment

Demultiplexing of the pooled samples and adapter removal was carried out using inbuilt algorithm of Minknow. Linux Long Time Support (v.20.04) Operating System was used for all the analysis. A comprehensive report of the sequencing was generated by NanoComp (https://github.com/NanoComp/h5utils) tool and sequencing quality assessment was done by FastQc tool (https://www.bioinformatics.babraham.ac.uk/projects/fastqc/). Sample reads were subjected to a minimum phred quality score of 9 and reads with lower quality were filtered out using NanoFilt (https://github.com/wdecoster/nanofilt) tool. Some initial reads are usually prone to low base-calling quality. Hence, 50bp of each initial reads from every sample were filtered out for quality maintenance using NanoFilt tool only. All the samples were subjected to quality assessment before and after quality filtering.

### Differential Gene Expression analysis

Differential Gene Expression analysis (DGE) was done using “pipeline-transcriptome-de” (https://github.com/nanoporetech/pipeline-transcriptome-de) pipeline. This pipeline from nanopore tech uses snakemake, minimap2, salmon, edgeR, DEXSeq and stageR to automate DGE workflows on long read data. Pipeline was set to make only reads aligned to minimum 3 samples to be considered for analysis. A separate conda (https://docs.conda.io/en/latest/#) environment was created on Linux OS to host this pipeline. Quantification and DGE was done by R language based (https://www.r-project.org/) tool edgeR (https://bioconductor.org/packages/release/bioc/html/edgeR.html) employing gene-wise negative binomial regression model and normalisation factor (Transcript Mean of M-value) for each sequence library. Differentially expressed genes (DEGs) with log2Fold Change (log2Fc) ≥0.5, ≤-0.5 and associated p-value <0.05 were selected as significant for further analysis. Annotation of DEGs was fetched from ENSEMBL database (https://asia.ensembl.org/index.html) using R language biomaRt package (https://bioconductor.org/packages/release/bioc/html/biomaRt.html). All the file compilation was ultimately done using Microsoft Excel and Libre Office calc.

Hierarchical clustering analyses was performed between all four samples of each group to generate Heatmap from normalised log2 counts per million expression values using web-based START(Shiny Transcriptome Analysis Resource Tool, https://kcvi.shinyapps.io/START/) tool. Comparative gene expression boxplot on the basis of log2 Counts per million between Mw and Control group were generated from START tool. Volcano plot involving all DEGs were created using R language based ggplot2 (https://ggplot2.tidyverse.org/) package.

### Gene Ontology (GO) Pathway Analysis

Pathway enrichment analysis was done for significant DEGs using g:Profiler (https://biit.cs.ut.ee/gprofiler/gost) using Gene Ontology. Pathways related to adaptive Natural killer (ANK), ADCC and innate immune inflammatory pathways were considered for focussed analysis. Functional enrichment was done by R based clusterProfiler (v. 4.2.1) https://bioconductor.org/packages/release/bioc/html/clusterProfiler.html tool. An adjusted p-value threshold of ≤0.05 was considered for this study.

### Statistics

Lymphocyte subsets have been represented as % of the parent population. Binary variables were compared between the groups using chi square test. The continuous variables were analysed using independent sample t-test considering Levene’s test for equality of variances and non-parametric tests (Mann-Whitney U test). P value < 0.05 was considered to be significant. GraphPad Prism (version 8.0 for Windows, GraphPad Software, La Jolla, CA, USA) was used for the statistical assessment (unpaired low-parametric Mann–Whitney or Kruskal–Wallis test and Spearman correlation). Recursive partitioning analysis was carried out using *rpart* package (https://cran.r-project.org/web/packages/rpart/index.html) in R (https://cran.r-project.org/) to generate optimal cut off for ANK cells at baseline. The efficacy of Mw in reducing the incidence of COVID-19 was calculated in terms of attack rates incidence risk ratio (IRR), absolute risk reduction (ARR) and intervention efficacy (see Table S3). This was calculated in terms of infections occurring at 2 weeks after administration of Mw is customary in prophylactic studies as well as those occurring any time between during the study period.

## RESULTS

### Mw and COVID-19

The characteristics and outcomes of the Mw and control groups are detailed in Table 1. All subjects were seropositive for CMV.

**Table 1:** Characteristics and Outcomes.

|  | Control group<br>(N=50) | Mw group<br>(N=50) | p-value |
| --- | --- | --- | --- |
| Age at vaccination, Median (Range), Years | 28 (21-55) | 28 (22-56) | 0.11 |
| <45 | 45 | 42 |  |
| >45 | 5 | 8 |  |
| Gender |  |  | 0.3 |
| Male | 34 | 28 |  |
| Female | 16 | 22 |  |
| SARS-CoV-2 Infection | 17 | 3 | 0.0012 |
| Mild | 11 | 3 | 0.02 |
| Moderate | 6 | 0 | 0.01 |
| Severe | 0 | 0 | 1.0 |
| Incidence rate/10000 person days | 24.05 | 3.57 |  |
| Time to Infection, Median (Range), Mean, Days | 56 (18-135)<br>66.4 ± 39.1 | 156 (151-159)<br>155.3 ± 4.0 | <0.0001 |

**Table 2:** Incidence Rate Ratio of SARS-CoV-2 infections and Mw efficacy.

| Parameter | Estimate | p-value | 95% CI |
| --- | --- | --- | --- |
| Incidence Rate Ratio (IRR) | 0.15 | 0.0004 | 0.04 to 0.5 |
| Attack rate in Mw control (ARU) | 0.34 |  |  |
| Attack rate in Mw treated (ARV) | 0.06 |  |  |
| Mw efficacy (%) | 82.35% |  | 50 to 96 % |
| Absolute Risk Reduction (ARR) | 0.28 |  | 0.13 to 0.43 |
| Number Needed to Treat (NNT) | 3.6 |  | 2.3 to 7.5 |

Twenty (3 in Mw group vs 17 in the control group) out of 100 subjects in the entire cohort had symptomatic COVID-19 during the study period (15 days-6 months). All infections on the Mw group were mild, while 6 of 17 in control group had moderate disease and required hospital admission (p=0.01). No deaths were recorded in either group. In Mw group all three infections occurred beyond 150 days. In the control group, 17 subjects had COVID-19 at a median of 56 days (range, 18–135). Additionally, three mild symptomatic COVID-19 were documented within the first 7 days of Mw administration. The incidence rates (IR) of COVID-19 were 3.57 and 24.05 /10,000 person-days in Mw and control groups respectively (IRR-0.15, 95% CI, 0.04-0.5, p=0.0004) with an efficacy rate for Mw being 82.35%, (95% CI, 50-96 %) (Fig. 1).

**Figure 1:**
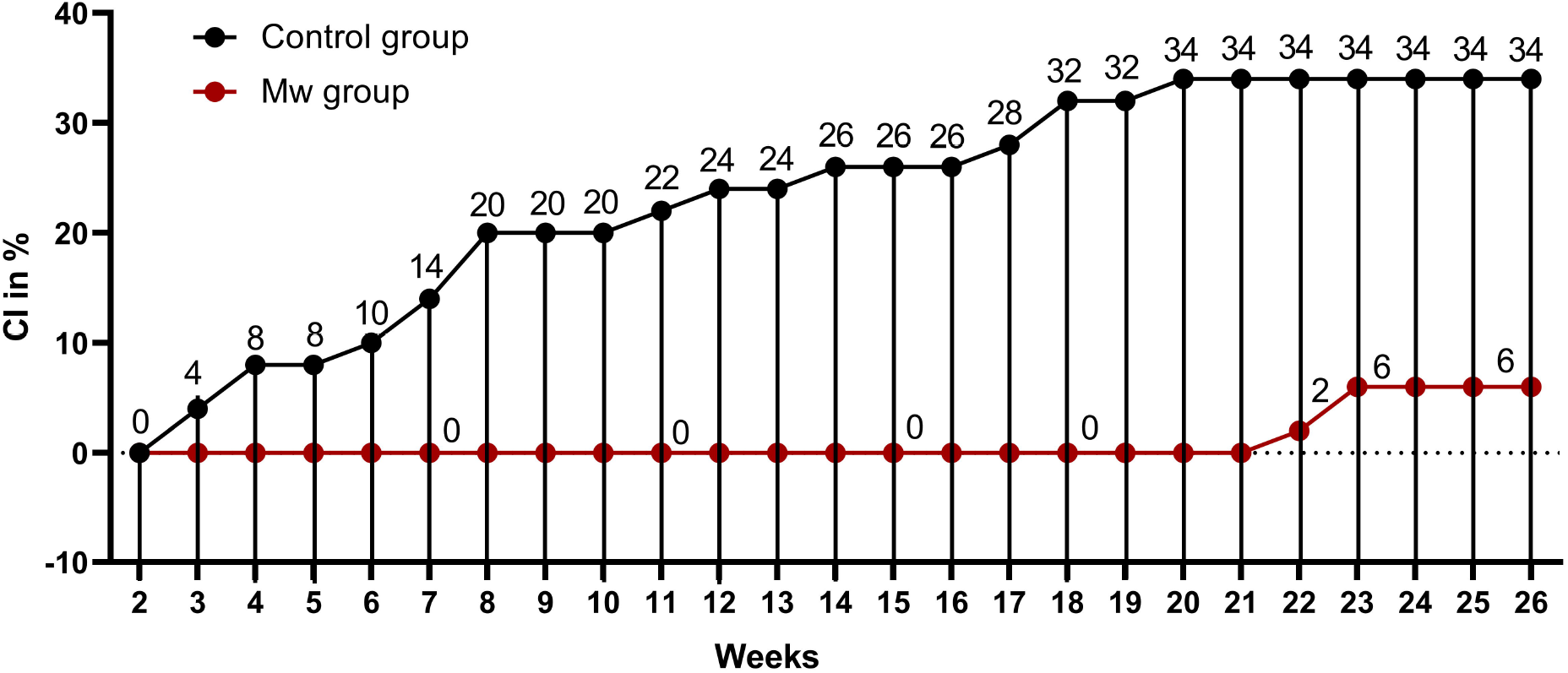
Impact of Mw vaccination on COVID-19 compared to a control group: Points and connecting line plot show the infection trend in Mw group (n=50) and the Control group (n=50). The x-axis shows the time in weeks and y-axis shows the Cumulative Incidence (CI) in %. Red and black shaded circles represent Mw group and Control group respectively.

Twenty-four subjects in the Mw group were randomised 1:1 to receive the second dose, 30 days after the first dose. Impact of the second dose on protection could not be analysed as none of the randomised subjects developed COVID-19 during the study period.

### Safety Profile of Mw

Mw was safe with no systemic adverse effects. Only one subject had mild fever (37.5° C) for less than two hours, 24 hours after administration, which was self-limiting. However, 14% had pain at the local sites, with 12% developing pain and erythema lasting for more than 72 hours. In 8% of the subjects, ulceration was noted at the local site with long-term scars similar to that observed with BCG vaccination. Those with severe local reactions were all older (35 years and above), compared to the median age of the group (28 years).

### Baseline NKG2C^+^ANK cells correlated with increased risk of COVID-19 in both Mw and Control groups

Baseline data on immune parameters was available in 80 subjects in the overall cohort, 30 in control and 50 in Mw group. The NKG2C^+^ANK cells in those who got infected (n=16) was 7.9 ± 6.1, (includes 3 who got infected within 7 days of receiving Mw) compared to 20.9 ± 13.8 in those who remained uninfected (p= 0.0005).

Amongst the 30 subjects in the control group, who were tested at baseline for NKG2C^+^ANK cells, 10 developed COVID-19. The NKG2C^+^ANK cells at baseline was 9.7 ± 6.6 in those with infection compared to 17.7 ± 5.56 in those without infection (p=0.0016). Those who acquired the infection in the Mw group, also had a lower baseline NKG2C^+^ANK (4.76 ± 3.9 vs 22.3 ± 16.1, Fig. 5, p=0.01). Interestingly, 2 of the 3 subjects, experiencing COVID-19 in the Mw group, did not show any increment in NKG2C^+^ANK levels or NKG2C/NKG2A ratios at any of the time-points. We did not find any correlation between expression of NKG2A as well as NKG2C/NKG2A ratio with development of symptomatic COVID-19 in this study.

### Mw and ANK cells

Thirty subjects from the Mw group and 15 from the control group, who did not develop COVID-19 or any other significant infection during the study period were included in the final longitudinal analysis of immunological impact of Mw.

### Baseline ANK levels did not differ, but ANK Kinetics at 60 days were different in the Mw group

There was no difference in overall NK cells between the two groups at baseline or at subsequent time-points. NKG2C^+^ANK and NKG2A^+^ iNK cells expression were also similar at baseline in both groups (Fig. 2, A & B). The data from control group shown in further comparisons only pertain to the baseline and day 60, as the values in the control group for both NKG2C^+^ ANK cells and NKG2A^+^ iNK cells did not change significantly at any of the time-points.

**Figure 2:**
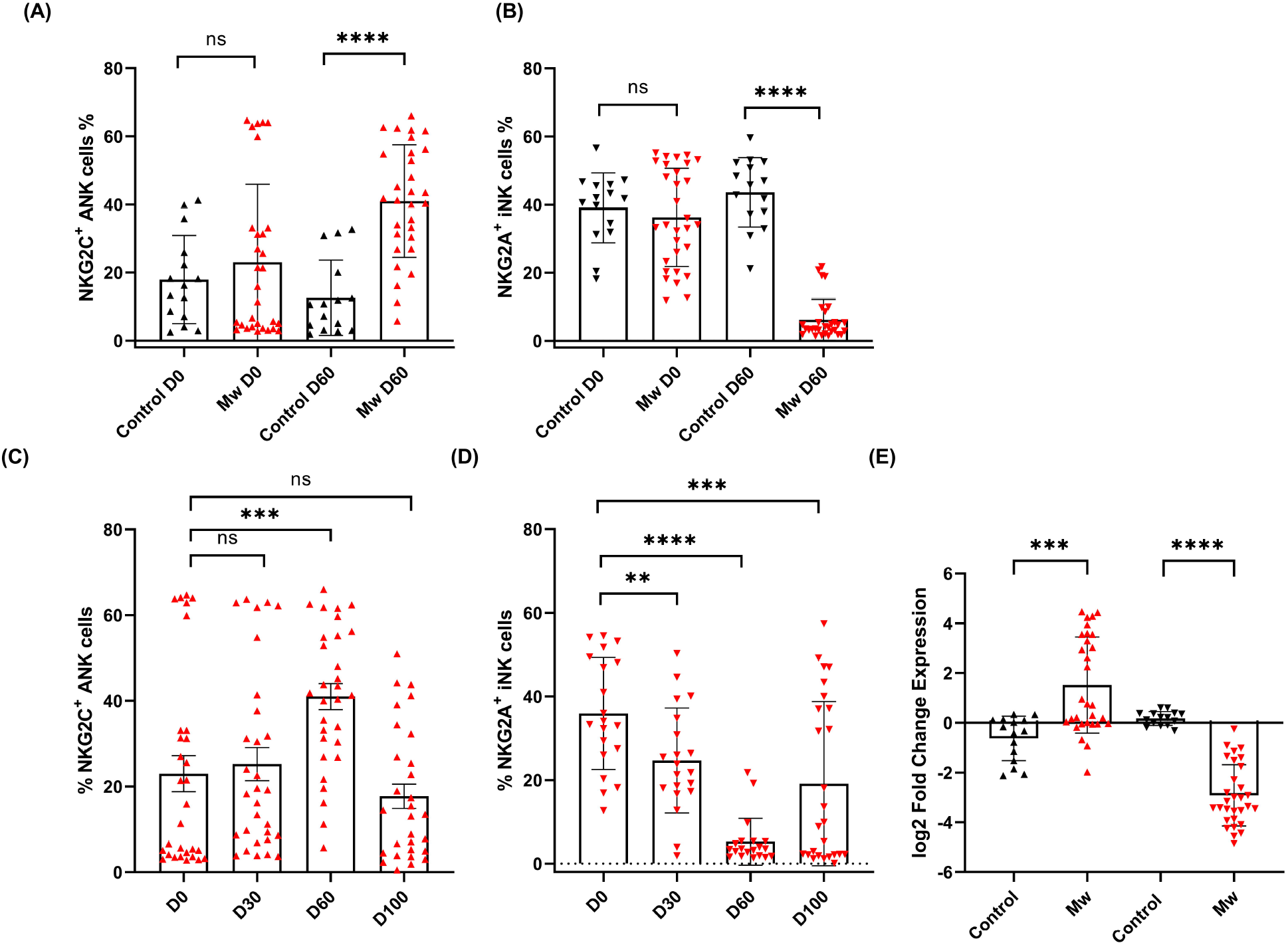
Impact of Mw on both Adaptive NK (ANK) cells and inhibitory NK (iNK) cells: A) Scatter dot with bar plot showing the expression of NKG2C^+^ ANK cells (Mw group - n=30 and control group- n=15) and B) NKG2A^+^ iNK cells (Mw group-n=30 and control group- n=15) with and without of Mw vaccine at baseline and day 60. C) Scatter dot with bar plot showing kinetics of NKG2C^+^ ANK cells, and D) NKG2A^+^ iNK cells expression in Mw group (n=30) at Baseline, day 30, day 60 and day 100. E) log2FC expression of NKG2C^+^ and NKG2A^+^ in both Mw group (n=30) and Control group (n=15) at day 60 after normalization with baseline. Red upside and downside shaded triangles represent NKG2C^+^ ANK and NKG2A^+^ iNK respectively for Mw group. Black upside and downside shaded triangles represent NKG2C^+^ ANK and NKG2A^+^ iNK respectively for Control group. ****- p < 0.0001, **- p <0.01, ***- p <0.001 and ns= p value not significant.

In the Mw group on the other hand, (Fig. 2, A-D), NKG2C^+^ANK cells increased from baseline (23.0 ± 22.9) to peak at day 60 (40.9 ± 16.5, day 60 p=0.0009, Fig. 2 C), while NKG2A^+^ iNK cells reduced from 36.23 ± 14.4 to 26.22 ± 14.04 on day 30 and 6.17 ± 6.01 on day 60 (p<0.0001, Fig. 2 D). The absolute values as well as the log two-fold change (log2FC) were significantly different for both NKG2C^+^ANK cells (1.5 ± 1.9 vs -0.62 ± 0.89, p=0.0002, Fig. 2 E) as well as NKG2A^+^ iNK cells (−2.9 ± 1.2 vs 0.17 ± 0.29, p<0.0001, Fig. 2 E) cells at day 60, compared to the control group.

### The impact of Mw on NKG2C upregulation was only observed in those with low ANK cells at baseline

A cut-off of 15% was established for NKG2C^+^ANK cells based on recursive partitioning for evaluating effect of base line value on NKG2C upregulation. Upregulation of NKG2C^+^ ANK cells was seen in those with NKG2C^+^ ANK levels below 15% (Low ANK group). From a baseline value of 4.7 ± 2.2, this increased to 8.9 ± 4.9 on day 30 (p=0.005,) and 43.3 ± 19.12 on day 60 (p<0.0001, Fig. 3 A). This was also reflected in the log2FC values.

**Figure 3:**
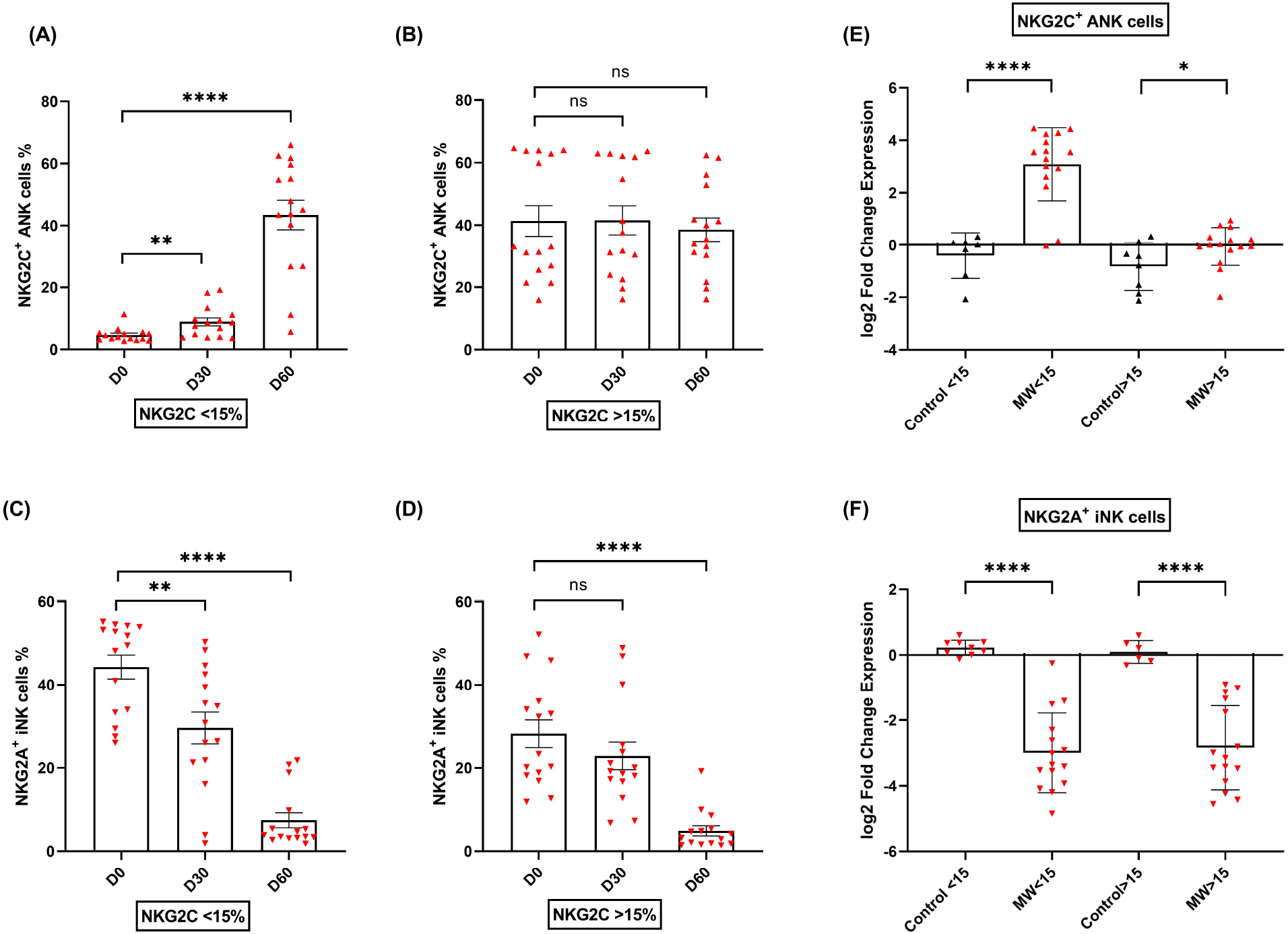
Impact of Mw on upregulation of NKG2C^+^ ANK cells with respect to NKG2C expression at baseline: Scatter dot with bar plot showing the kinetics of A) NKG2C^+^ ANK cells (n=15) and C) NKG2A^+^ iNK (n=15) cells expression in Mw group at baseline, day 30 and 60 in respect to <15% NKG2C at baseline. B) NKG2C^+^ ANK (n=15) cells and D) NKG2A^+^ iNK (n=15) cells expression in Mw group at baseline, day 30 and 60 in respect to >15% NKG2C at baseline. Log2FC expression of E) NKG2C^+^ ANK and F) NKG2A^+^ iNK cells at day 60 after normalization with baseline value in Mw group (>/<15 %, n=15) and control group. Red upside and downside shaded triangles represent NKG2C^+^ ANK and NKG2A^+^ iNK respectively for Mw group. Black upside and downside shaded triangles represent NKG2C^+^ ANK and NKG2A^+^ iNK respectively for Control group. ****- p < 0.0001, **- p < 0.01, *- p < 0.05 and ns= p value not significant.

In those with NKG2C^+^ANK levels above 15% (High NKG2C^+^ANK), there was no significant change in absolute values of NKG2C^+^ANK cells at either day 30 or day 60(Fig. 3 B), but a positive impact on log2FC was noted (p=0.04, Fig. 3 E).

### The impact on NKG2A^+^ iNK downregulation was irrespective of baseline NKG2C^+^ANK expression

The downregulation of NKG2A^+^iNK cells was noted in the Mw group, irrespective of baseline NKG2C expression [Fig. 3, C & D]. The downregulation of NKG2A^+^iNK cells was significant at day 60 in the Mw group, irrespective of baseline NKG2C expression (−2.99 ± 1.21 vs 0.23 ± 0.23, p<0.0001 for Low ANK group and -2.84 ± 1.28 vs 0.09 ± 0.35, p<0.0001 for High ANK group) [Fig. 3 F].

### Mw had a sustained effect on NKG2C+ANK/ NKG2A+iNK ratio until 100 days

NKG2C^+^ ANK/NKG2A^+^ iNK ratio was similar between two groups at baseline. However, this increased in the Mw group at day 60 (12.3 ± 9.2, p<0.0001) and persisted until day 100 (7.44 ± 13.55, p=0.01). This was witnessed for both Low and High ANK group in the Mw cohort but not in the control cohort. [Fig. 4, A-D].

**Figure 4:**
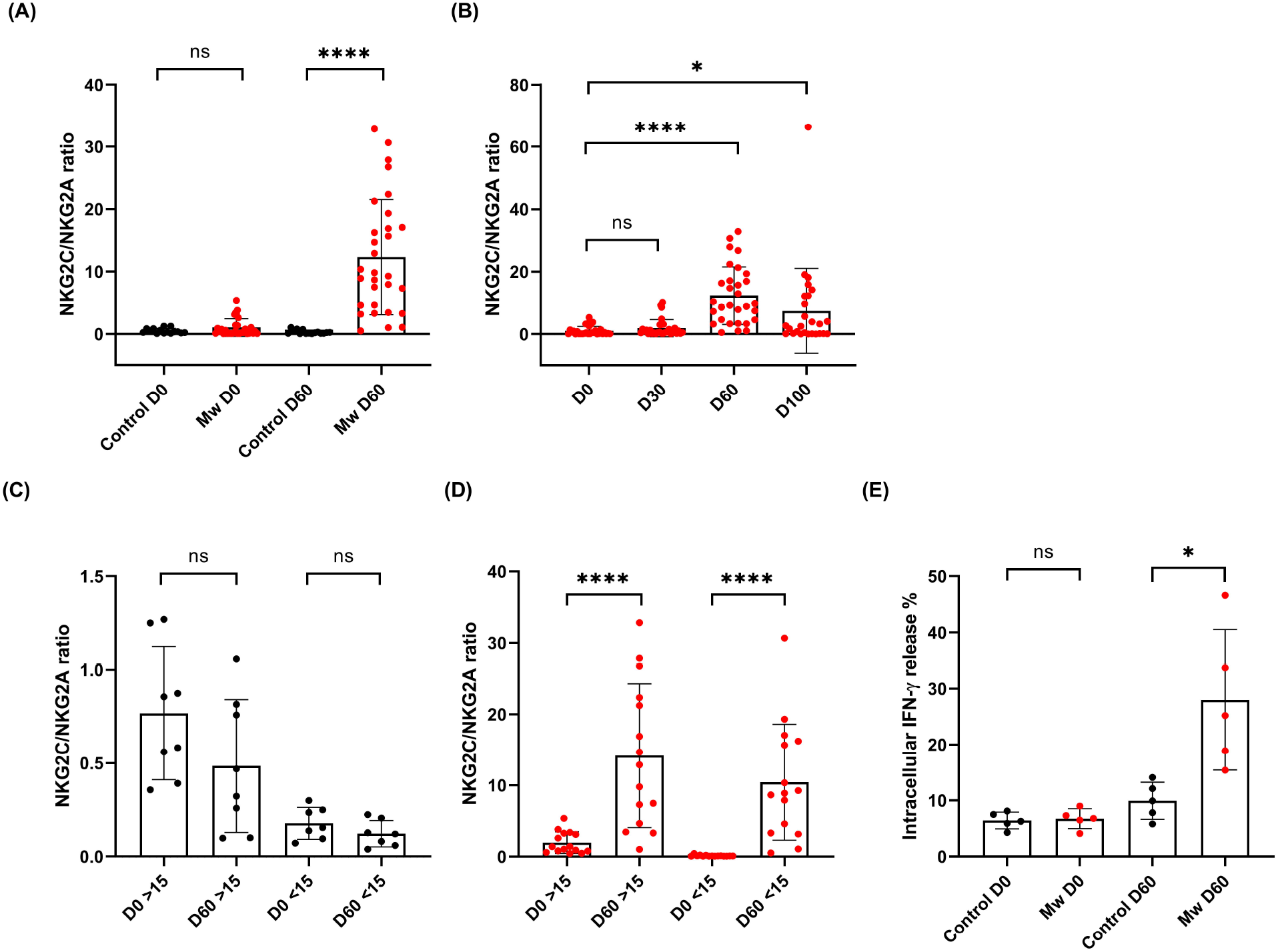
Mw showed sustained effect on NKG2C/NKG2A ratios until 100 days and Impact of Mw intracellular Cytokine (IFN-γ) release: Scatter dot with bar plot showing A) NKG2C/NKG2A ratio of Control group (n=15) and Mw group (n=30) at baseline and day 60. B) Kinetics of NKG2C/NKG2A ratio at baseline, day 30, 60 and 100 in Mw group. C) & D) Kinetics of NKG2C/NKG2A ratio at baseline and day 60 on the basis of >/< 15 % NKG2C at baseline in Mw group and control group respectively. E) intracellular IFN-γ release in both control (n=5) and Mw (n=5) group at baseline and day 60. Red and black shaded circles represent Mw group and Control group respectively. ****- p < 0.0001, ***- p < 0.001, *- p < 0.05 and ns= p value not significant.

### IFN-γ release was higher in Mw group at 60 days

IFN-γ release potential of the NKG2C^+^ANK cells was studied at baseline and at 60 days. This was similar between the groups at the baseline (mean-6.7 vs 6.4). However, this was significantly increased in the Mw group at day 60, compared to the control group (mean-27.96 vs 9.9, p=0.01, Fig. 4 E).

### KLRC2 and kinetics of ANK cells

KLRC2 deletion was studied in the Mw group only. KLRC2 *Wt* ^*+*^*/del*^*+*^ was detected in 36% of the subjects in the Mw group. This was not associated with any significant decrease in the baseline NKG2C^+^ANK levels (15.1 ± 17.67 vs 20.4 ± 17.96, p=0.46). There was no difference in the effect of Mw between the KLRC2 *Wt*^*+*^*/ Wt*^+^ and W*t*^*+*^*/Del*^*+*^ groups at day 60 either (p=0.26), although the log2FC increase tended to be higher in the W*t*^*+*^*/Del*^*+*^ group (p=0.12) [Fig. S1, A & B]. KLRC2 deletion was detected in 2 out of 6 with COVID-19, compared to 16 out of 44 without COVID-19 (p=0.6).

### Mw did not influence kinetics of naïve and memory T cell subsets

CD4 and CD8 cells remained unchanged, as were the CD45RA and CD45RO subsets at days 30 and 60 in the Mw group [Fig. S2, A & B]. The same was witnessed in the control group (data not shown).

NKG2A expression on CD3^+^ T cells was examined at baseline and subsequent timelines. Despite a significant downregulation of NKG2A observed in NK cells, no such effect was evident in the T cells. [Fig. S2 C]

### Impact of the second dose of Mw on NKG2C^+^ANK cells

There was no difference between in NKG2C^+^ANK cells at day 60 and day 100 (p=0.28) in double vs single doses of Mw (Fig. 6 A). However, NKG2A^+^ iNK cell expression was significantly reduced at day 100 (7.7 ± 16.77 vs 24.85 ± 19.43, p=0.04) in double dose group (Fig. 6 B). The NKG2C/NKG2A ratio was also significantly higher in double dose group at day 100 (10.42 ± 7.19 vs 2.08 ± 3.26, p <0.001) [Fig. 6 C]. There was no difference in CD4^+^ & CD8^+^ T cells and memory or naïve subsets at day 60 and 100 between the randomized groups (data not shown).

**Figure 5:**
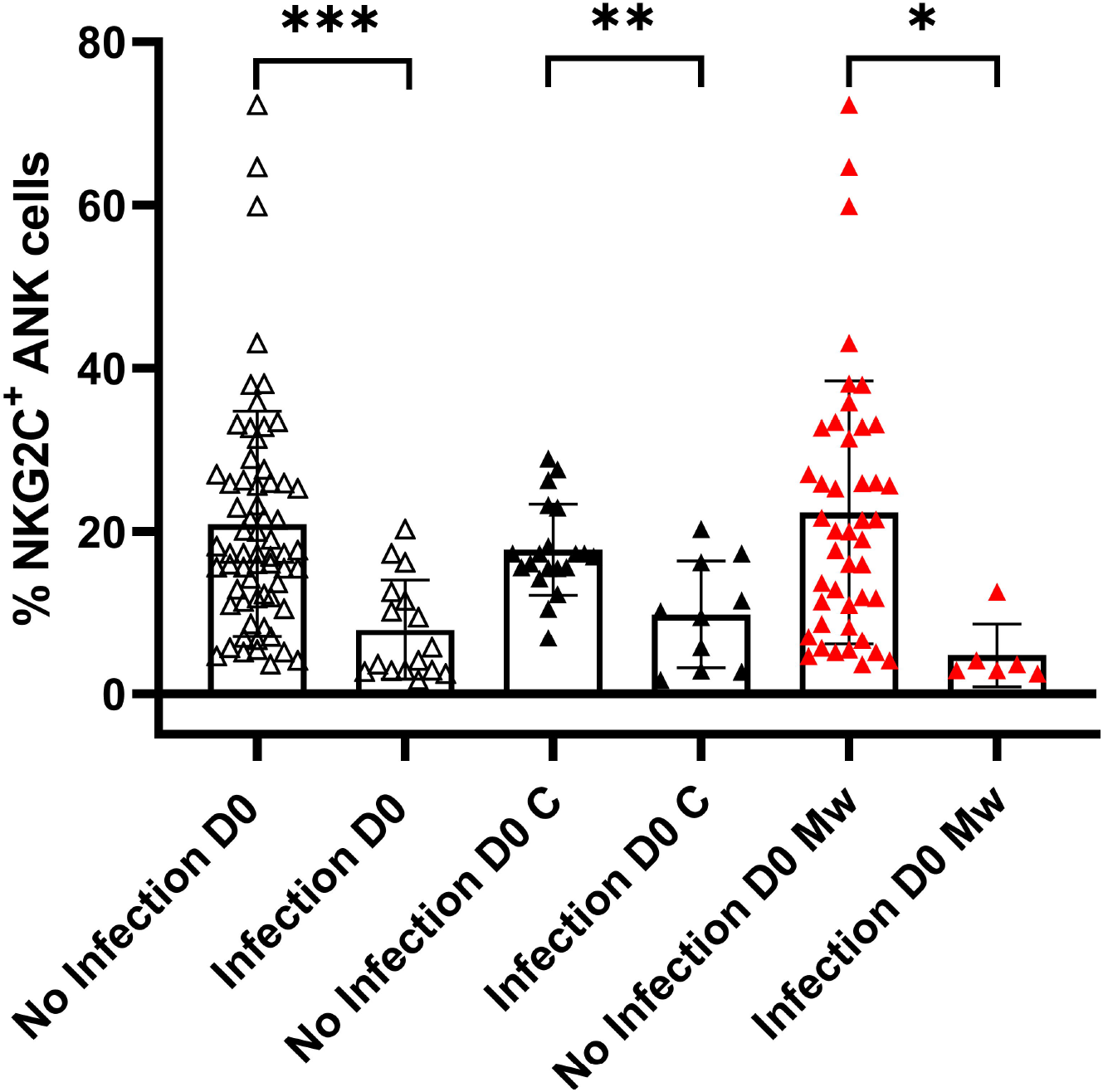
Relationship between NKG2C^+^ANK cells and COVID-19: Scatter dot with bar plot showing expression of NKG2C^+^ ANK cells at baseline in the overall cohort (no infection, n=64 & infection, n=16), Control group (no infection, n=20 & infection, n=10) and Mw group (no infection, n=44 & infection, n=6) with respect to SARS-CoV-2 infection. Black and red upside shaded triangles represent NKG2C^+^ ANK for Control group and Mw group respectively and unshaded upside triangles represent NKG2C^+^ ANK of both groups combined. ***- p < 0.001, **- p < 0.01 and * - *P* < 0.05.

**Figure 6:**
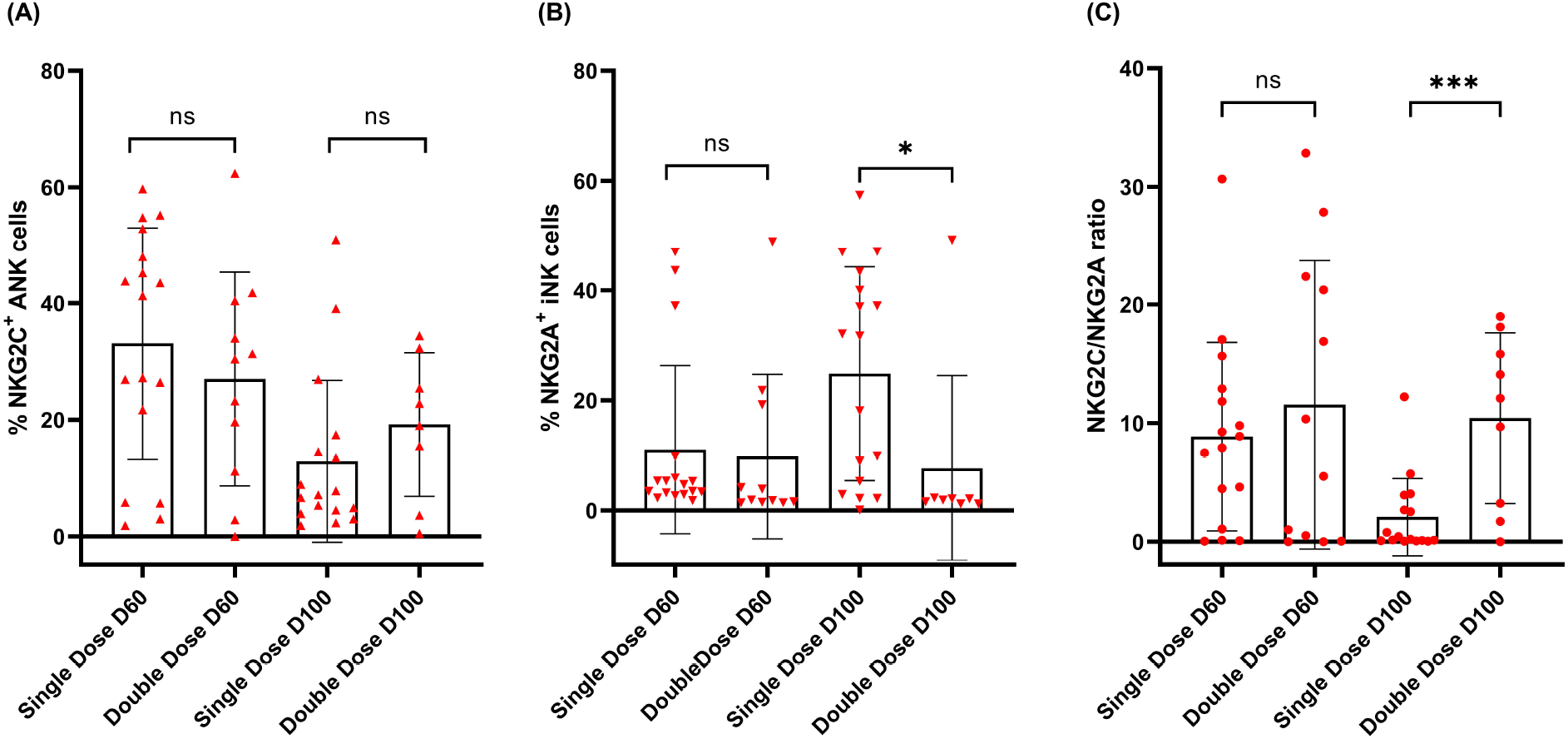
Impact of the Second dose of Mw on Adaptive and Inhibitory NK cells: Scatter dot with bar plot showing expression of A) NKG2C^+^ ANK (day 60, single dose; n=17, double dose; n=12 and day 100, single dose; n=17, double dose; n=12), B) NKG2A^+^ iNK cells (day 60, single dose; n=12, double dose; n=12 and day 100, single dose; n=17, double dose; n=12) and C) NKG2C/NKG2A (day 60, single dose; n=17, double dose; n=12 and day 100, single dose; n=17, double dose; n=12) ratio with respect to single and double dose of Mw vaccine at day 60 and 100. Red upside and downside shaded triangles represent NKG2C^+^ ANK and NKG2A^+^ iNK respectively for Mw group. Red shaded circle represents Mw group. ****- p < 0.0001, **- p < 0.01, *- p < 0.05 and ns= p value not significant.

### RNAseq Analysis at 6 months

Sequencing and mapping metrics along with Quality scores are shown in Supplement 1 (Table S1)

### Identification of differentially expressed genes

Based on criteria of p-value < 0.05, a total of 3,603 out of 16,058 were found to be differentially expressed genes (DEGs) between Mw and Control group, including 1568 genes that were upregulated and 2035 genes that were downregulated in Mw group. Those DEGs with a log2 fold change of at least 0.5 were considered for further analysis. DEGs related to Adaptive Natural Killer Cells and ADCC were selected out to analyse the differential expression pattern between two groups.

### ANK and ADCC genes were upregulated in Mw group

Nineteen DEGs were associated with ANK and ADCC pathways (Fig. 7, A & B). 11 genes were upregulated and 8 were found to be downregulated in Mw group. KLRC2 (NKG2C), BCL11B, ARID5B, B3GAT1 (CD57), KLRC4 were upregulated and KLRC1(NKG2A), ZBTB16 (PLZF1), KIT and SH2D1B (EAT-2) were downregulated in relation to ANK pathway (Fig. 7 C). In the ADCC pathway, CD247 (CD3ζ), FCGR1A (CD64), FCGR2A(CD32a), FCGR2C (CD32c) were upregulated and FCER1G was downregulated (Fig. 7 C), favouring ANK mediated ADCC.

**Figure 7:**
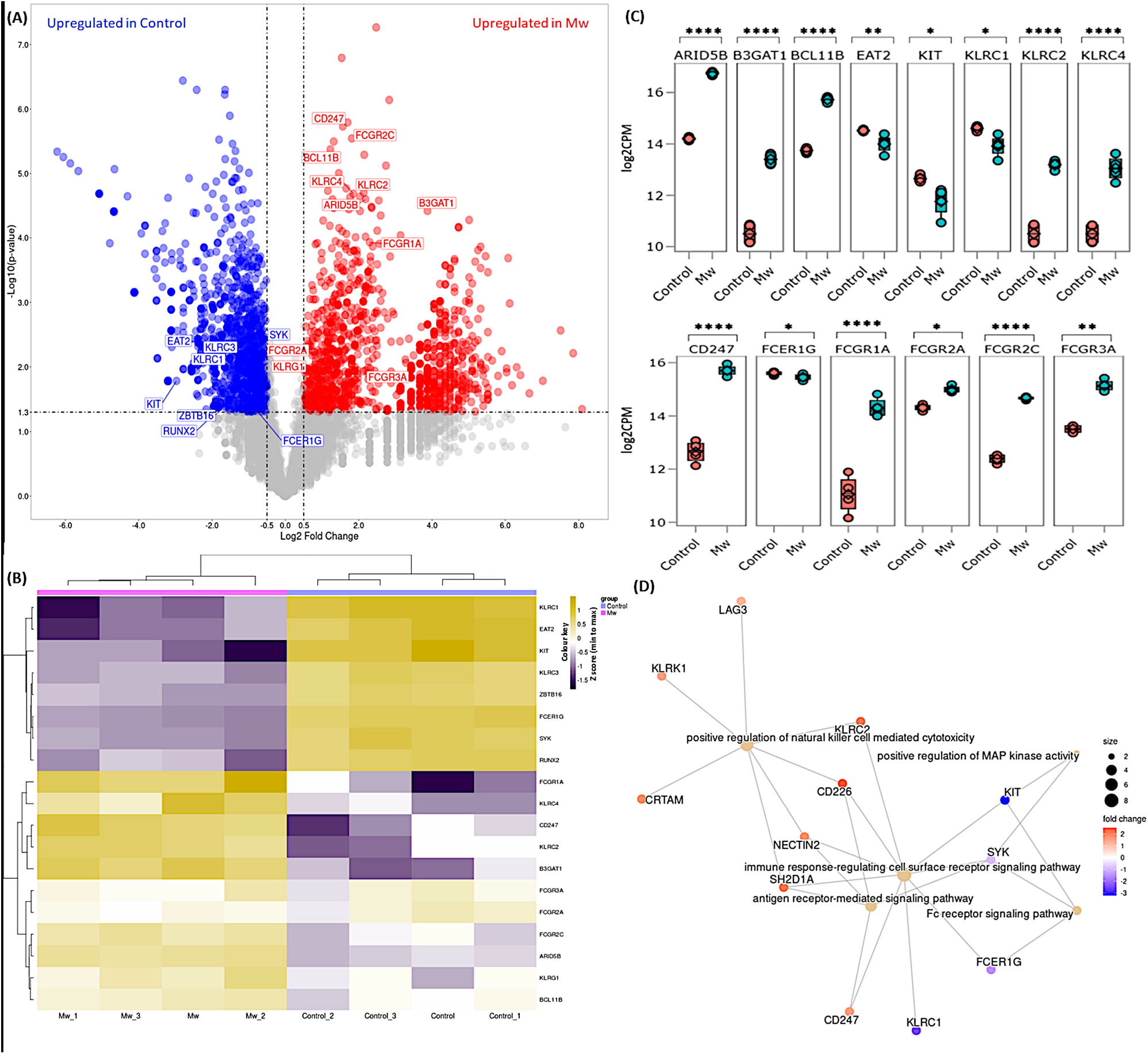
Differential gene expression and network analysis in Mw and Control group: A) Volcano plot showing genes differentially expressed in Mw as compared to control group. Genes with a p-value <0.05 and log2 Fold Change >=0.5 <= -0.5 were considered significant. Selected genes are highlighted (Red: Upregulated in Mw group, Blue: Upregulated in Control group). B) Heatmap of RNA-seq expression data showing selected DEGs related to this study. Hierarchical cluster analysis were performed between all samples of Mw and Control group. Gene expression is shown in normalized log2 counts per million. C) Boxplots showing expression levels of 19 selected genes in Mw (green) as compared to control group (orange). Relative gene expression is shown in normalized log2 counts per million (log2CPM) for I) ANK pathway in the top panel, II) ADCC pathway in the middle panel. (*p-value < 0.05, **p-value < 0.01, ***p-value < 0.001, ****p-value<0.0001). D) GO network analysis of the top 5 enriched GO terms in the differentially expressed genes for Adaptive NK cell and Antibody-dependent Cellular Cytotoxicity (Fisher’s exact test using enrichGO function in R package clusterProfiler, multiple test correction by Benjamini-Hochberg method, adj. p-value <0.05).

### Gene Ontology (GO) Pathway Analysis

Functional enrichment of differentially expressed genes were carried out on the selected genes focusing on ANK and ADCC pathways. Selected 19 DEGs were analysed using goProfiler and clusterProfiler for Gene Ontology (GO). GO analysis of the selected DEGs demonstrated that immune response-regulating cell surface receptor signalling pathway (GO:0002768) and positive regulation of natural killer cells mediated toxicity (GO:0045954) were significantly upregulated (Fig. 7 D).

## DISCUSSION

The identification of NK cells with ability to persist and mount recall responses has challenged the traditional compartmentalization of adaptive and immune responses[22; 23]. The NKG2C^+^ANK cells explored in this study are known to be induced by primary exposure to CMV[24; 25]. However, in CMV exposed patients, these NKG2C^+^ANK cells have been found to proliferate in response to a variety of viral pathogens, including HIV, HCV, Hantavirus, influenza, and pox viruses[16; 26; 27; 28; 29]. The unique nature of this non-antigen specific recall response against a wide variety of pathogens, puts NKG2C^+^ANK cells at the forefront in the battle against novel pathogens such as SARS-CoV-2 [3; 30; 31; 32; 33; 34], where antigen-specific adaptive response generated from T and B cells are absent at the outset of the pandemic.

BCG vaccine has been shown to induce long-term heterologous memory response of NK cells[31]. This has been studied, both in the context of infections as well as cancer[35]. Influenza vaccination has been shown to induce proliferation of NKG2C^+^ANK cells with potential for enhanced cytokine release[36]. These considerations prompted us to explore this novel immunomodulator, Mw, which has been extensively studied as adjuvant or prophylaxis in other infections[19; 37; 38], and some forms of cancers including BCG resistant bladder cancer [39].

Our study demonstrated that prophylaxis with Mw was associated with a six-fold reduction in the incidence of symptomatic COVID-19 in this high-risk cohort over a 6-month period. This protective efficacy was accompanied by a sharp increase in NKG2C^+^ANK cells between 30- and 60-days following exposure to Mw. A critical observation was a steep decline in the expression of the inhibitory counterpart, NKG2A^+^iNK cells, which continued through 100 days. This resulted in an increasing NKG2C/NKG2A ratio through the study period. Exposure to a second dose of Mw resulted in a continued downregulation of NKG2A^+^ iNK cell and a further increase in NKG2C/NKG2A ratios. The importance of the relative proportion of NKG2C and NKG2A expressing NK cells could be appreciated better in the context of the fact that the inhibitory impact of NKG2A overrides that of NKG2C, as it binds to the ligand HLA-E with several fold greater affinity than NKG2C[5]. In the context of SARS-CoV-2 infection, in-vitro studies have demonstrated that the viral spike protein-1 (SP-1) upregulates NKG2A on NK cells and HLA-E on the infected lung epithelial cells, causing a strong inhibition of cytotoxicity of NK cells[40]. Our group has demonstrated a correlation between high expression of NKG2A, suppression of NKG2C, and adverse outcome following severe COVID-19 lung disease, despite viral clearance[34]. It is possible that the adverse effect of KLRC2 deletion genotype as reported earlier[30], was mitigated by Mw. Hence, it might be inferred that if any intervention can achieve the reverse, i.e., downregulation of NKG2A and upregulation of NKG2C expression, this might offer innate protection against COVID-19.

In our study, a lower baseline NKG2C^+^ANK cells predisposed to COVID-19 in the control group. The same in the Mw group was associated with COVID-19 in the first 2 weeks, but not thereafter until 150 days. It is also worth noting that COVID-19 occurring in the Mw group beyond 150 days was associated with failure in modulation of the adverse NKG2C^+^ANK cell profile in 2 out of 3 subjects. Apart from phenotypic alteration, Mw seemed to influence the cytokine release potential as well, as evidenced by an increase in IFN-γ release at day 60 in the Mw group, compared to both baseline as well as the control group. Furthermore, upregulation of NKG2C was dominantly noted in those with a lower baseline NKG2C^+^ANK cell. Upregulation of NKG2C expression might be more tightly controlled, unlike downregulation of NKG2A, in mature and licensed NK cells, where an inhibitory regulation mediated by NKG2A is physiologically redundant due to KIR-mediated inhibitory control. More importantly, this also demonstrates that downregulation of checkpoint receptor NKG2A is an important determinant of a favorable ANK profile.

RNA-seq analysis confirmed that upregulation of ANK pathway was evident at 6 months in the Mw group. Apart from upregulation of KLRC2 and B3GAT1 and downregulation of KLRC1, the key transcription factor in the ANK pathway, BCL11b, was persistently upregulated[41]. Downregulation of EAT-2 and PLZF further corroborated the classic gene expression signature of ANK cells. Moreover, increased expression of AT-rich interaction domain 5B (ARID5B), as demonstrated in the Mw group plays an important role in enhanced metabolism in ANK cells as well as increased IFN-γ release and survival[42]. DGE analysis also revealed an enhancement of ANK mediated ADCC pathway, with significant upregulation of CD247 along with downregulation of FCER1G, which is a typical signature of ANK-ADCC[43]. Both CD247 and FCER1G are adapter molecules for FCGRIIIA (CD16) with CD247 possessing 3 ITAMs against one ITAM of FCER1G, increasing the ADCC several folds[44]. It is possible that Mw induced augmentation of NK-ADCC might potentiate the efficacy of SARS-CoV2 vaccines as well[45]. Prior studies on Mw have explored its effect on TLRs via the monocyte/macrophage pathway[46; 47; 48] and not on NK cells. The suggested mechanistic pathway as to how Mw might be favorably influencing ANK mediated protection against COVID-19 has been depicted in Fig. 8.

**Figure 8:**
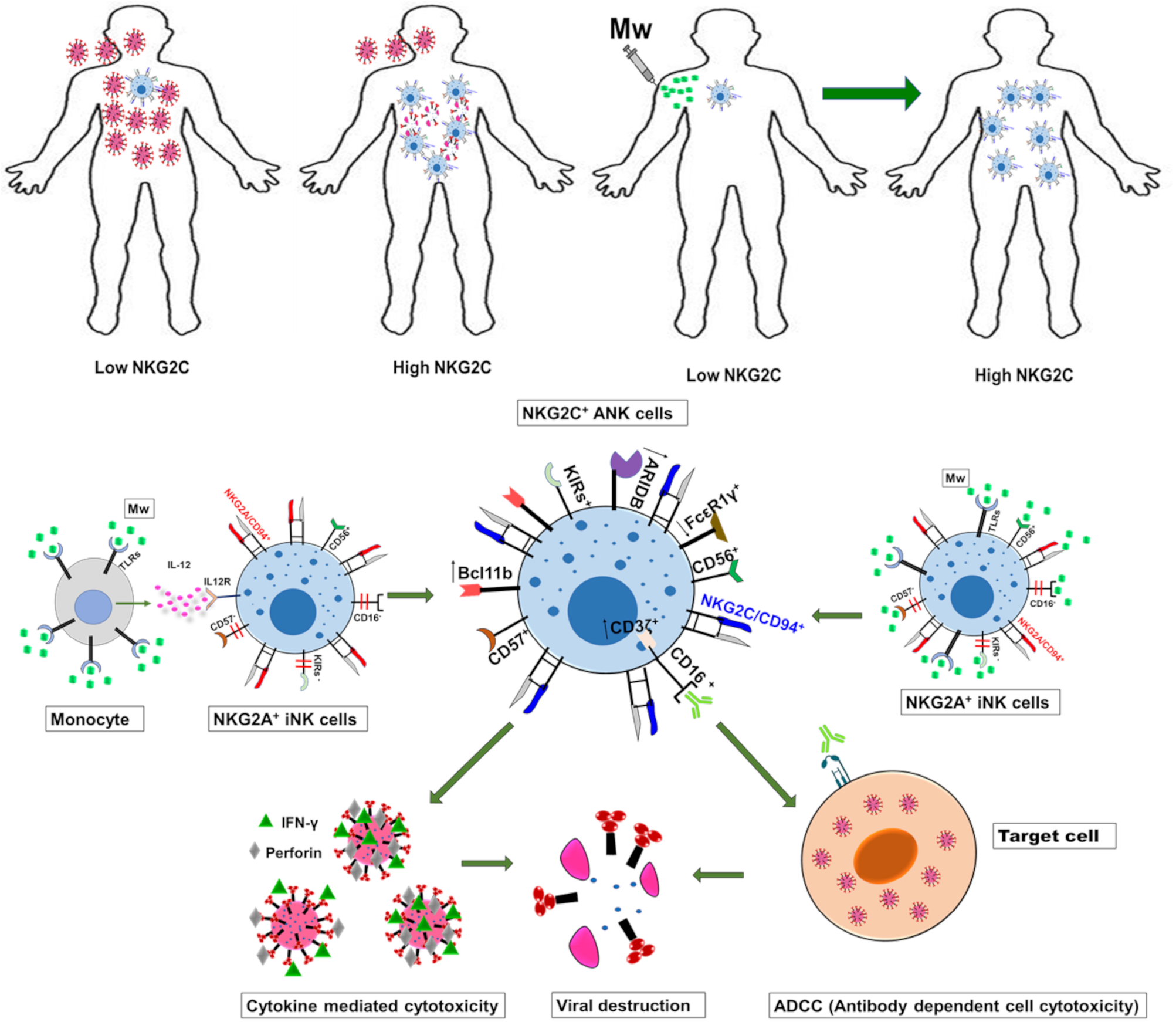
The suggested mechanistic pathway of the impact of Mw on ANK cells and protection against COVID-19.

In conclusion, our study shows that Mw offered protection against COVID-19 in a high-risk population at the peak of the pandemic in the absence of antigen-specific immunity. This could be through a sustained upregulation of NKG2C^+^ANK cell and ADCC pathway, along with simultaneous downregulation of NKG2A^+^iNK pathways. If borne out in a randomized setting, these findings might usher a novel approach to bolster heterologous immunity in the current and future pandemics and identify ANK pathway in the immunological vulnerability for developing COVID-19.

## Data Availability

All data produced in the present work are contained in the manuscript.

## Acknowledgement

We thank all the nursing and technical staff of the department who have assisted in carrying out this study. This study was supported by a grant from Indo-US Science and Technology Forum (IUSSTF/VN-COVID/049/2020).

## Authorship Contribution

SRJ, BK and SC designed the study. AM, RL, GB and HM performed the study. SRJ, AS, AM, GB and HM collected the data. SRJ, AS, JA, SVM, DT and SC analyzed the data; SRJ and SC wrote the manuscript. All the co-authors reviewed and approved the manuscript.

## Data Availability Statement

The RNA-seq data generated from the experiment related to study have been uploaded in ‘Sequence Read Archive’ (SRA)

