## Supplemental Figures 1-5S for "Impact Of an Immune Modulator Mycobacterium-w On Adaptive Natural Killer Cells and Protection Against COVID-19"

**Figure S1:**

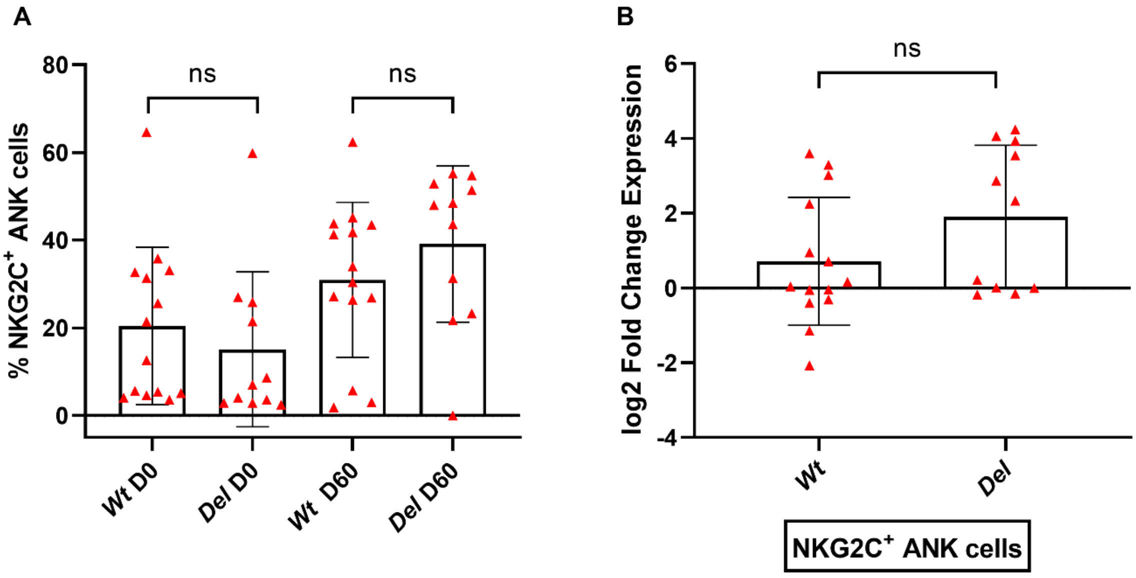


**Figure S1: KLRC2 genotyping does not impact the effect of Mw on NKG2C$^{+}$ ANK cells:** Scatter dot with bar plot showing (A) Expression of NKG2C$^{+}$ ANK cells with respect to KLRC2 genotyping (WT- wild type and DEL-Deletion) at baseline (WT, n=14, DEL, n=11) and day 60 (WT, n=14, DEL, n=11). (B) log2FC expression of NKG2C$^{+}$ ANK cells at day 60 after normalization with baseline NKG2C$^{+}$ expression comparison of KLRC2 WT (n=14) and DEL (n=11) genotyping. Red upside shaded triangles represent NKG2C$^{+}$ ANK for Mw group. ns= P value not significant.

**Figure S2:**

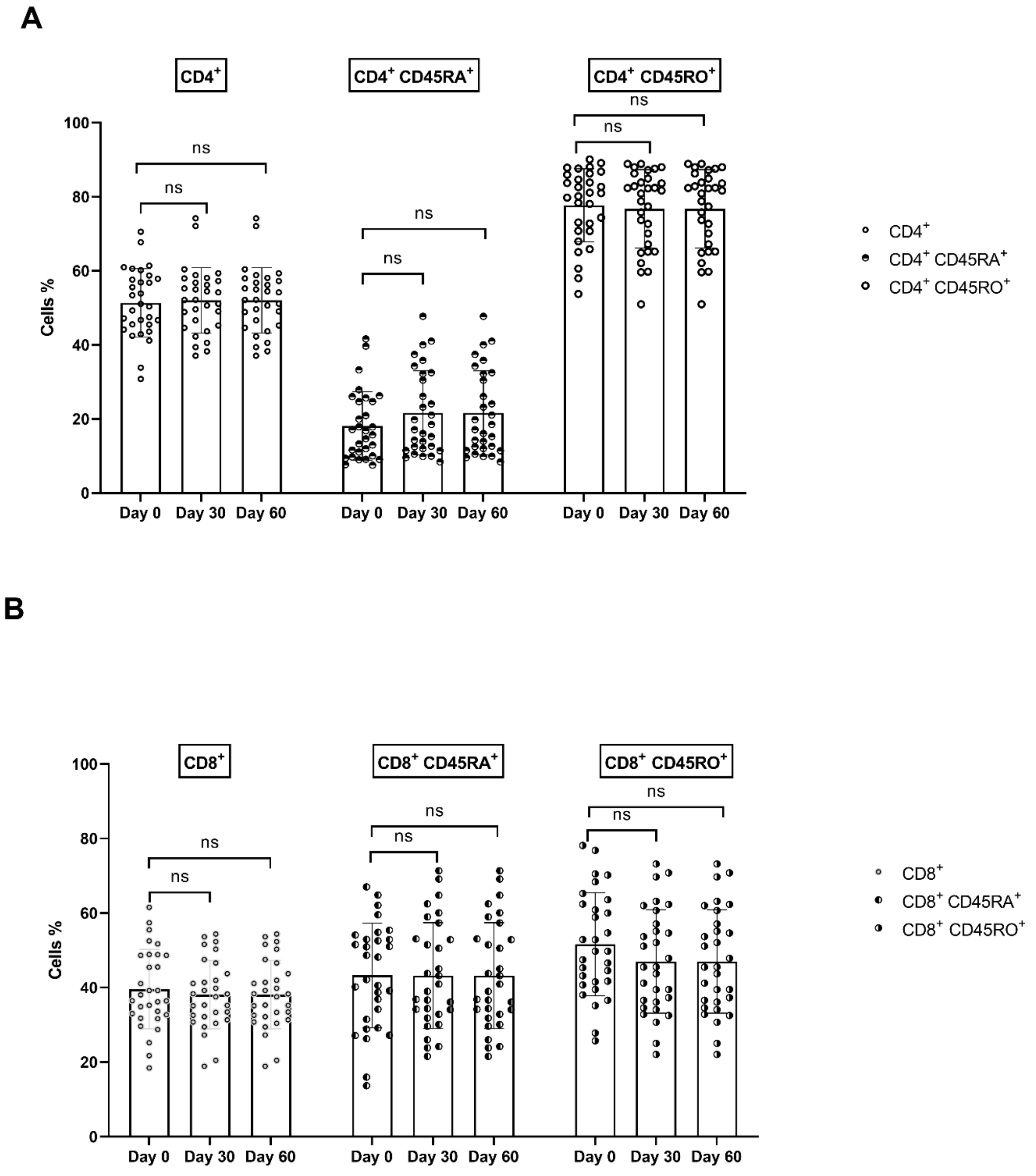
A
CD4+
CD4+ CD45RA+
CD4+ CD45RO+
Cells %
100
80
60
40
20
0
ns
ns
ns
ns
ns
ns
Day 0
Day 30
Day 60
Day 0
Day 30
Day 60
Day 0
Day 30
Day 60
CD4+
CD4+ CD45RA+
CD4+ CD45RO+
B
CD8+
CD8+ CD45RA+
CD8+ CD45RO+
Cells %
100
80
60
40
20
0
ns
ns
ns
ns
ns
ns
Day 0
Day 30
Day 60
Day 0
Day 30
Day 60
Day 0
Day 30
Day 60
CD8+
CD8+ CD45RA+
CD8+ CD45RO+

C

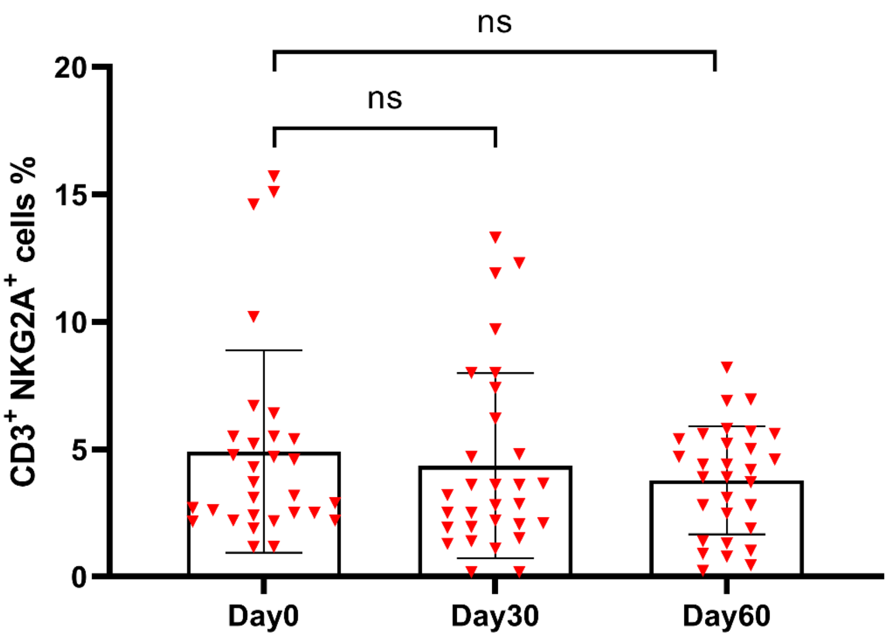


**Figure S2: Mw does not affect the T cell subsets $CD4^+$, $CD8^+$ and $CD3^+$ $NKG2A^+$:** Scatter dot with bar plot showing at baseline, day 30 and day 60: (A) Kinetics expression of $CD4^+$, $CD4^+CD45RA^+$ and $CD4^+CD45RO^+$, (n=30), (B) $CD8^+$, $CD8^+CD45RA^+$, and $CD8^+CD45RO^+$ (n=30). (C) Scatter dot with bar plot showing at baseline, day 30 and day 60: $CD3^+$ $NKG2A^+$ (n=30) T-cell subsets. $CD4^+$(◦), $CD4^+CD45RA^+$(◓) and $CD4^+$ $CD45RO^+$(○), $CD8^+$(⨀), $CD8^+CD45RA^+$(◐), and $CD8^+CD45RO^+$ (◑). Red downside shaded triangles represent $CD3^+$ $NKG2A^+$ for Mw group. ns= P value no significant.

**Figure S3:**

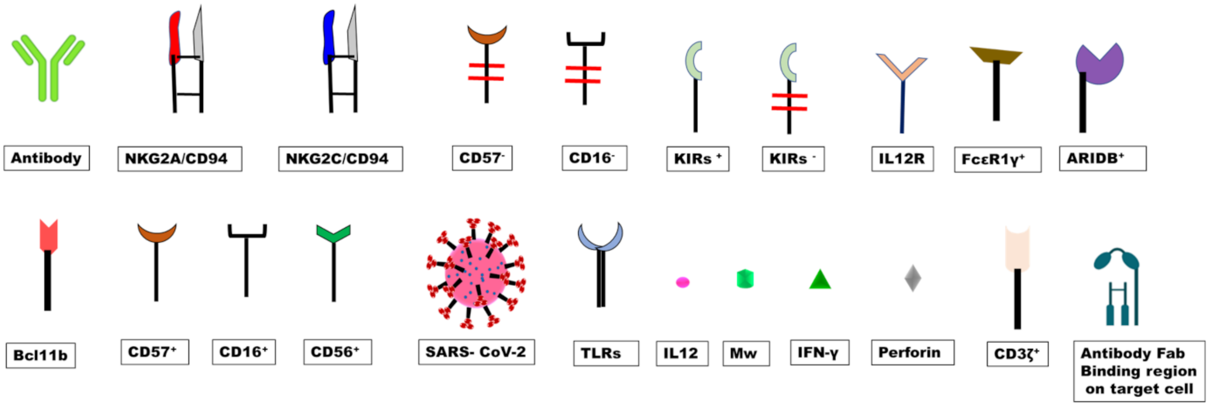


**Figure S3:** Key to the cartoon depicted in **Figure 8**

**Figure S4:**

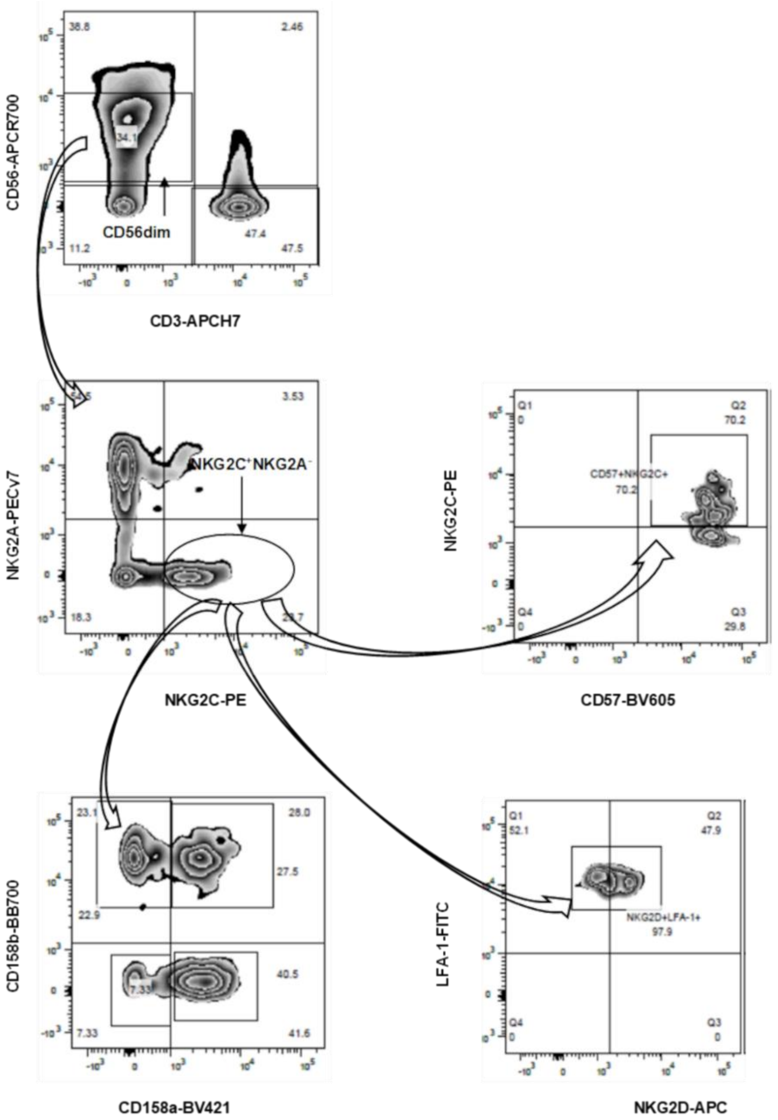


**Figure S4 Flow cytometry gating strategy for NK cells:** PBMC were stained for NK cell markers CD56, CD57, NKG2C, NKG2A, NKG2D, CD158a, Cd158b and T cell marker CD3. (Top left panel) Shows the gating of NK cell from lymphocytes by selecting CD56$^{+}$ CD3$^{-}$ population. (Middle left panel) shows the gating of NKG2C$^{+}$/NKG2A$^{-}$ and NKG2C$^{-}$/NKG2A$^{+}$ population gated from CD56$^{+}$ CD3$^{-}$ NK cells. (Middle right panel) shows the gating of NKG2C$^{+}$CD57$^{+}$ NK cell population from NKG2C$^{+}$/NKG2A$^{-}$. (Bottom left panel) shows the CD158a and CD158b NK cell population gated from NKG2C$^{+}$/NKG2A$^{-}$. (Bottom right panel) shows the gating of LFA-1 and NKG2D population gated from NKG2C$^{+}$/NKG2A$^{-}$.

**Figure S5:**

A B C

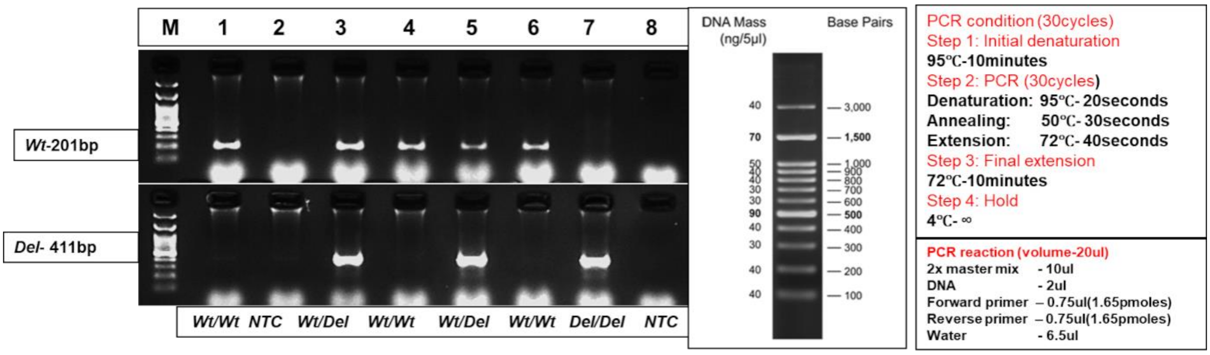


**Figure S5: KLRC2 genotyping by conventional PCR**: (A)Above image represents the KLRC2 Conventional PCR gel image. DNA samples were isolated from the human samples. Isolated DNA samples were used for the PCR amplification of KLRC2 Wild type(*Wt*) and Deletion (*Del)* gene (Primers details were mentioned in the methodology). For conventional PCR, 2µl of DNA samples were used and 30 cycles were used for amplification for both conditions. Top gel image shows the Wild type results and bottom image shows the Deletion type results. Samples labelled as M- 100bp DNA ladder, Samples 1- Homozygous Wild type positive control, Sample 2- NTC(non template control), Sample 3- Heterozygous both Wild type and Deletion positive control, Sample 4- Homozygous Wild type positive , Sample 5- Heterozygous both Wild type and Deletion positive, Sample 6- Homozygous Wild type positive, Sample 7- Homozygous Deletion positive Sample 8- NTC (Non Template Control). (B) 100bp DNA ladder data sheet image. (C) PCR condition and PCR reaction preparation
