## Supplemental table 1-3 for "Impact Of an Immune Modulator Mycobacterium-w On Adaptive Natural Killer Cells and Protection Against COVID-19"

**Supplemental Tables:**

**Table S1:**

**<u>RNAseq Analysis</u>**
**Sequencing and mapping metrics**

Average base calling phred quality score (q) was found to be more than 11 for each sample (range 8-30). Read base quality ≥ 9 was considered for DGE analysis. N50 of read length for sequenced pooled samples was more than 1kb (range 1kb-1.4kb). Average read-length was more than 800bp for each sample (range 200bp-4.6kb). Numbers of reads were more than 1 million for each sample. Total number of passed (>7 q score) base sequenced for each sample was found to be more than 700 million with one sample going as much as 1 billion implying a coverage of 21X to 35X. More than 60% mapping coverage was found for each sample. (See, Table S1)

**Sequencing and mapping metrics of samples (pass reads)**

| Sample ID | Mapping Percentage | Number of reads | Mean read length (nt) | Average base quality score (q) |
|---|---|---|---|---|
| NB04_1 (Control) | 67.20 | 1,582,130 | 868.4 | 11.2 |
| NB04_2 (Control) | 69.30 | 1,611,117 | 801.9 | 11.3 |
| NB05_1 (Control) | 66.76 | 1,561,297 | 946.2 | 11.3 |
| NB05_2 (Control) | 70.87 | 1,818,543 | 809.6 | 11.4 |
| NB01 (Mw) | 63.57 | 1,270,259 | 932.7 | 11.2 |
| NB02 (Mw) | 66.42 | 1,443,542 | 850.1 | 11.2 |
| NB03 (Mw) | 70.24 | 2,106,378 | 871.8 | 11.1 |
| NB06 (Mw) | 72.12 | 2,150,894 | 1039.8 | 11.3 |

**Table S2:**

| S.No. | Primers | Sequences |
|---|---|---|
| 1. | KLRC2 *Wt* Forward primer | *5' CAGTGTGGATCTTCAATG 3'* |
| 2. | KLRC2 *Wt* Reverse primer | *5' TTTAGTAATTGTGTGCATCCTA 3'* |
| 3. | KLRC2 *Del* Forward primer | *5' ACTCGGATTTCTATTTGATGC 3'* |
| 4. | KLRC2 *Del* Reverse primer | *5'ACAAGTGATGTATAAGAAAAAG 3'* |

**Table S3:**

**Statistical Method for calculation of efficacy of Mw**

| | Infected | Non-infected | Total |
|---|---|---|---|
| Mw treated | a | b | n1 |
| Mw control | c | d | n2 |

Attack rate in Mw control (ARU) = c/n2

Attack rate in Mw treated (ARV) = a/n1

Incidence Risk Ratio (IRR)/ Relative Risk = ARV/ARU

Absolute Risk Reduction (ARR) = ARU- ARV

Number needed to treat (NNT) =1/ARR

Vaccine Efficacy (%) = ((ARU-ARV)/ARU)*100

95% CI Vaccine efficacy (%) = 1- IRR CI
